# Year-round RSV Transmission in the Netherlands Following the COVID-19 Pandemic - A Prospective Nationwide Observational and Modeling Study

**DOI:** 10.1101/2022.11.10.22282132

**Authors:** Yvette N. Löwensteyn, Zhe Zheng, Neele Rave, Michiel A.G.E. Bannier, Marie-Noëlle Billard, Jean-Sebastien Casalegno, Virginia E. Pitzer, Joanne G. Wildenbeest, Daniel M. Weinberger, Louis Bont, the SPREAD study group

**Affiliations:** Department of Pediatric Immunology and Infectious Diseases, Wilhelmina Children’s Hospital, P.O. Box 85090, 3508 AB Utrecht, the Netherlands; Department of Epidemiology of Microbial Diseases and the Public Health Modeling Unit, Yale School of Public Health, New Haven, CT, USA; Department of Pediatric Respiratory Medicine, Maastricht University Medical Center, Maastricht, the Netherlands; Department of Microbiology, Hospices Civils de Lyon, Lyon University Medical Center, Lyon, France; Admiraal de Ruyter Ziekenhuis; Albert Schweitzer Ziekenhuis; Alrijne Ziekenhuis; Amsterdam Universitair Medisch Centrum; St. Antonius Ziekenhuis; Antonius Zorggroep Sneek; Beatrix Ziekenhuis; Bernhoven Ziekenhuis; BovenIJ Ziekenhuis; Bravis Ziekenhuis; Catharina Ziekenhuis; Dijklander Ziekenhuis; Elizabeth-TweeSteden Ziekenhuis; Elkerliek Ziekenhuis; Erasmus Medisch Centrum; Flevoziekenhuis; Franciscus Gasthuis & Vlietland; Gelre Ziekenhuis; Groene Hart Ziekenhuis; Haaglanden Medisch Centrum; IJsselland Ziekenhuis; Isala; Leids Universitair Medisch Centrum; Maastricht University Medical Center; Martini Ziekenhuis; Maxima Medisch Centrum; Meander Medisch Centrum; Medisch Centrum Leeuwarden; Noordwest Ziekenhuisgroep; Onze Lieve Vrouw Gasthuis; Radboud Universitair Medisch Centrum; Rijnstate Ziekenhuis; Slingeland Ziekenhuis; Spaarne Gasthuis; Tergooi Medisch Centrum; Treant Zorggroep; Universitair Medisch Centrum Groningen; VieCuri; Wilhelmina Ziekenhuis Assen; Zaans Medisch Centrum; Ziekenhuis Rivierenland; Ziekenhuis St Jansdal; Ziekenhuis Tjongerschans; ZorgSaam; Zuyderland Medisch Centrum; Rijksinstituut voor Volksgezondheid en Milieu

**Keywords:** Respiratory syncytial virus, transmission, epidemic timing, seasonality, COVID-19, waning immunity, immunity debt

## Abstract

A nationwide prospective study showed year-round RSV transmission in the Netherlands after an initial 2021 summer outbreak. The pattern was unprecedented and distinct from neighboring countries. Our dynamic simulation model suggests that this transmission pattern could be associated with waning immunity because of low RSV circulation during the COVID-19 pandemic.

## INTRODUCTION

Before the COVID-19 pandemic, RSV epidemics occurred annually during winter in temperate climates (**Figure 1A**). During winter 2020/2021, RSV was virtually absent following the implementation of COVID-19-related non-pharmaceutical interventions (NPIs) [1, 2]. After NPIs relaxed, various patterns of re-emergent RSV epidemics were observed in different countries. Several studies suggested that school reopening is associated with increased RSV activity [1, 3]. However, little attention has been paid to the impact of increased RSV susceptibility in children and adults due to low RSV exposure during the pandemic (“immunity debt”) on the patterns of resurgence [4].

**Figure 1.**
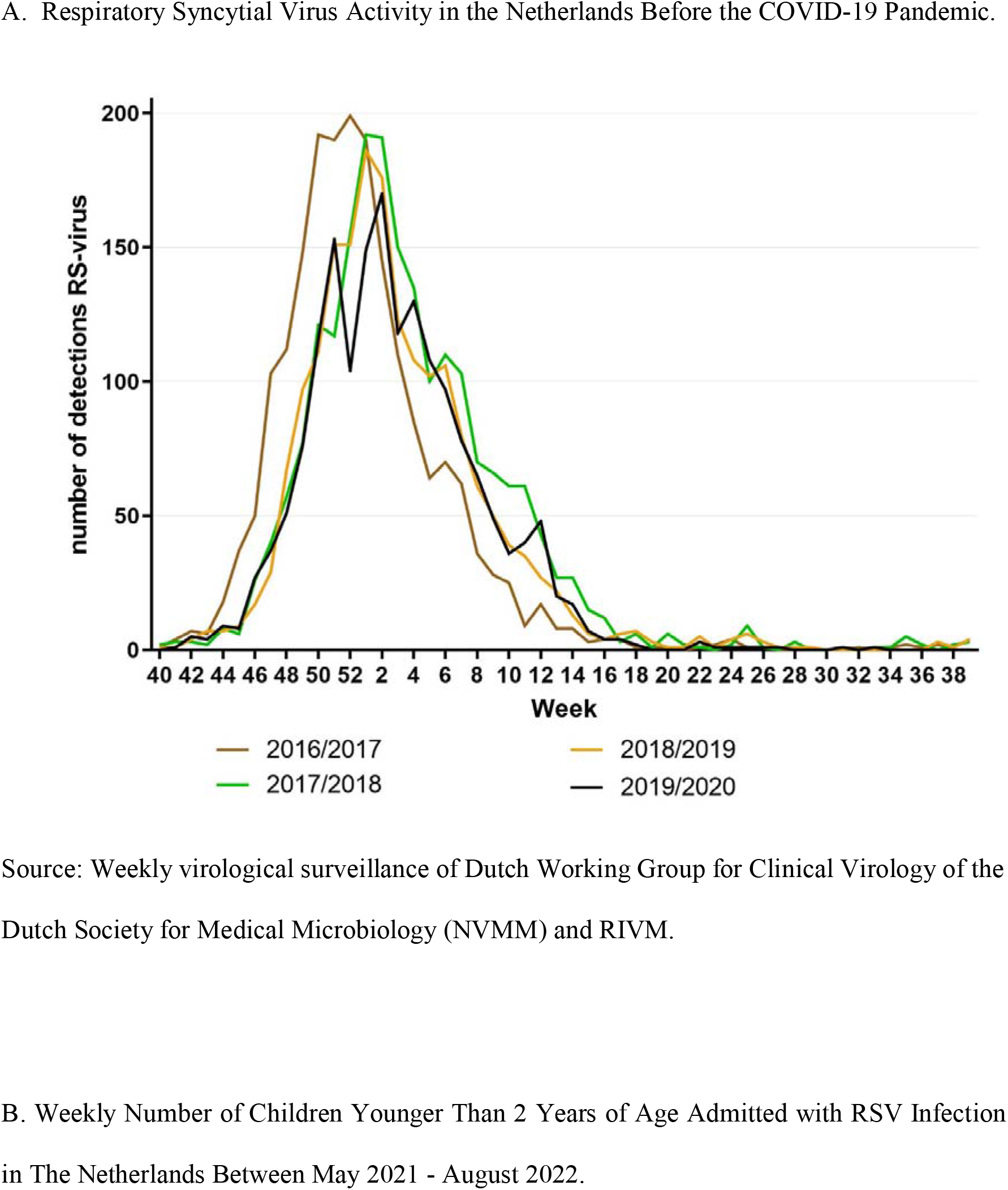
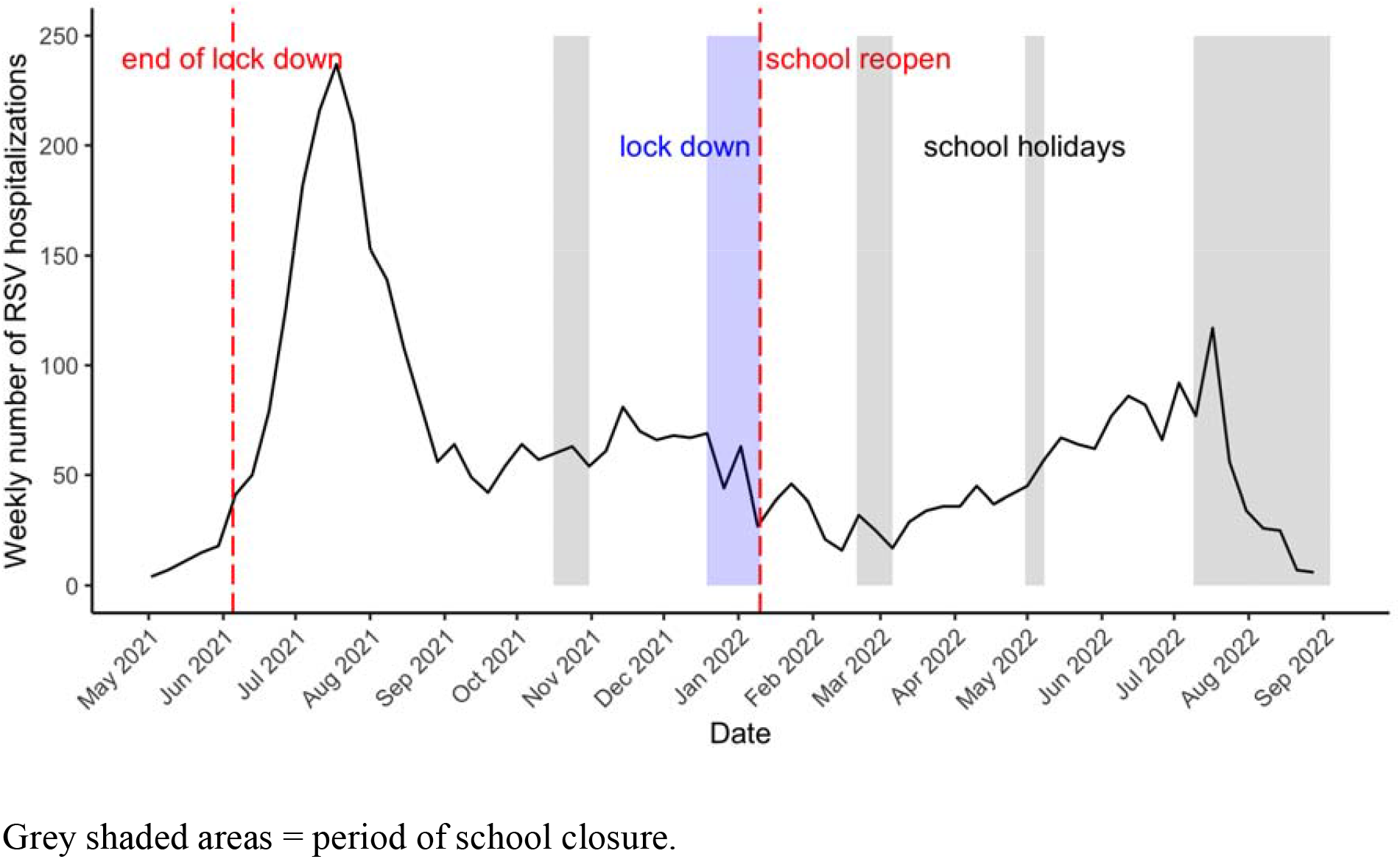
Respiratory Syncytial Virus Activity in the Netherlands.

Using a combination of prospective surveillance data from the Netherlands and simulation models, this study aims to (1) describe the unusual endemic pattern of RSV between May 2021 and August 2022 and (2) illustrate the impact of waning population immunity on the timing, intensity, and persistence of the re-emergent RSV epidemic. This study provides an opportunity to advance our understanding of the drivers of re-emergent RSV epidemics.

## METHODS

### Study Design

We initiated a prospective nationwide surveillance study (SPREAD study: surveillance of pediatric respiratory admissions in Dutch hospitals) in which real-time data are collected on RSV-related pediatric hospitalizations in 46 hospitals throughout the Netherlands (**Supplementary Table 1**). In 10 hospitals with standard-of-care RSV testing, clinical patient data are collected (**Supplementary Figure 1**). For this paper, we collected data from May 3, 2021 (week 18) until September 4, 2022 (week 35). Age-stratified data were collected from October 2018 until June 2022. The study population included all children between 0-2 years of age who were hospitalized with RSV-bronchiolitis in participating hospitals. Collaborators of participating hospitals were requested to share data on a weekly basis.

Follow-ups and data verification were performed by the study team to ensure data quality. Data were entered into the Castor Electronic Data Capture (EDC) system [5]. We analyzed potential differences in the age distribution of patients during the following periods: pre-COVID-19 (2018/2019 and 2019/2020 winter season (October-April)), post-COVID-19 summer outbreak (May-August 2021) and post-COVID-19 endemic phase (September 2021-August 2022). Retrospective data were collected using diagnosis treatment combination (DBC) codes (3210 - RSV-bronchiolitis; 3208 - lower respiratory tract infection; 3104 - upper respiratory tract infection), and RSV-positive admissions were manually selected using patient files. We used the Mann-Whitney-U test to determine statistically significant differences (defined as p<0.05) between subgroups. Analyses were performed with SPSS version 26.0 (IBM Corp, Armonk, NY).

### Evaluating Different Hypotheses Using Simulation Models

We modified our previously published age-stratified RSV transmission model that accounts for population dynamics, RSV seasonality, and virus importation from external sources to account for waning immunity (**Supplementary Modeling Methodology**) [6]. Our previous model assumed that individuals gain partial immunity following infection, which reduces their susceptibility to subsequent infections. By adding waning immunity, we assumed that individuals can become susceptible to infection after a long period of low virus exposure.

Several factors could explain differences in the timing and intensity of RSV epidemics before and after the COVID-19 pandemic, including length and strength of NPIs, RSV importation from external sources, increased birth rate in the Netherlands during COVID-19, and waning population immunity against RSV due to absent RSV circulation. We simulated RSV transmission from July 2018 to June 2025 to evaluate the various factors’ impact on the projected trajectories of RSV hospitalizations and qualitatively compared these to the observed re-emergent RSV epidemic in the Netherlands. The four scenarios that we evaluated were (1) moderate level of NPIs and low level of virus importation, no waning immunity; (2) moderate level of NPIs and low level of virus importation, with waning immunity; (3) moderate level of NPIs and high level of virus importation, with waning immunity; (4) strict NPIs but high level of virus importation, with waning immunity (**Supplementary Table 5**). We developed a free-to-use web-based Shiny app to allow researchers to simulate re-emergent RSV under the impact of various factors. We provide an interactive example at https://3wxpl3-zhe-zheng.shinyapps.io/shiny/. Data and code to reproduce this study are available from GitHub (https://github.com/weinbergerlab/SPREAD.git).

## RESULTS

### RSV-related Pediatric Hospitalizations

Starting from May 24, 2021 (week 21), a summer outbreak of RSV was observed. During the peak week (July 19, week 29), 240 patients were admitted with RSV-bronchiolitis. Subsequently, continuous RSV transmission was observed, with RSV-related admissions stabilizing at approximately 50 patients weekly (**Figure 1B**).

### Older Patients During and After the 2021 Summer Outbreak

We obtained age-stratified data for 988 patients admitted with RSV infection for 2018-2022 in 10 hospitals (**Supplementary Table 2**). The proportion of patients <6 months was higher during pre-COVID-19 winter seasons. Median age during pre-COVID-19 winter seasons 2018/2019 and 2019/2020 was 69 days (IQR 35-176). During the summer outbreak, median age increased to 161 days (IQR 56-383) (p<0.001). During the endemic phase (September 2021 - August 2022), median age decreased again to 137 days (IQR 54-281) but was still higher than during pre-COVID winter seasons (p<0.001) (**Supplementary Figure 2**). As a sensitivity analysis, we excluded patients from the only academic hospital (UMCU), which contributed a large number of patients (21% pre-COVID-19 versus 11% during the summer outbreak and 14% during the endemic phase). Differences between groups remained unchanged (**Supplementary Table 3**).

### Association of Waning Population Immunity with RSV Epidemic Timing

A model that assumed moderate NPIs, low virus importation, and waning population immunity against RSV due to low RSV circulation (Scenario 2) most closely resembled the Dutch situation: a large summer outbreak followed by continuous RSV transmission (**Figure 2**). Under this scenario, the proportion of RSV hospitalizations in children >1 year was expected to be higher during the summer outbreak than during a typical winter season. This proportion decreased over time during the following “endemic” phase. Alternative scenarios that assumed no waning RSV immunity failed to generate a summer outbreak in 2021 that was more intense than previous winter epidemics (**Supplementary Figure 4A**). A model that assumed strict NPIs, high level of virus importation, and waning immunity after a prolonged period of low viral exposure (Scenario 4) most closely resembled RSV activity in Germany and France: RSV activities returned to normal winter epidemics after a small outbreak in Spring 2021 (**Supplementary Figure 4C**).

**Figure 2.**
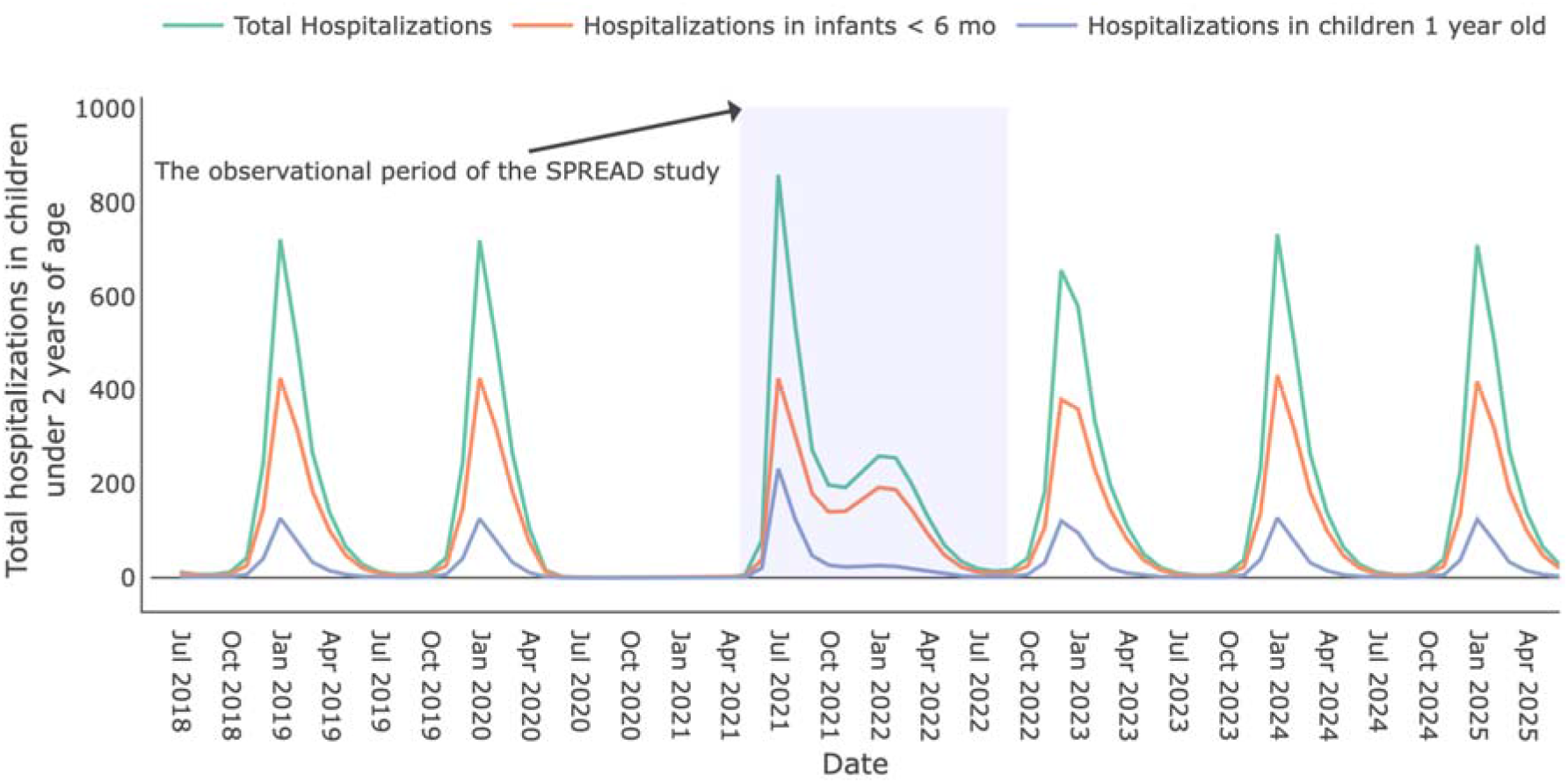
Simulated number of monthly respiratory syncytial virus (RSV) hospitalizations in children <2 years of age in the Netherlands, 2018–2025, for the model including waning of immunity. The year 2018-2019 corresponds to the RSV hospitalizations before the COVID-19 pandemic. We assumed that COVID-19-related public health measures started in April 2020 and gradually relaxed since March 2021 over a 3-month period [11]. The projected RSV hospitalizations after the interruption of COVID-19-related measures over time are plotted for 2021 to 2025. The green line corresponds to the total RSV hospitalizations in children under 2 years of age. The orange line corresponds to the RSV hospitalizations in infants under 6 months of age. The purple line corresponds to the RSV hospitalizations in children 1 year of age. The shaded area corresponds to the period of the SPREAD study, May 2021 - August 2022.

## DISCUSSION

The Netherlands exhibited a unique pattern of RSV re-emergence during the COVID-19 pandemic characterized by a high summer peak in 2021 followed by a prolonged period of continuous transmission at mid- to high-level RSV activity. Our model simulations confirm that population “immunity debt” can explain the large RSV summer outbreak and the following “endemic phase” (**Supplementary Figure 5**) [4, 7-9]. Additionally, NPIs were gradually re-implemented between mid-November 2021 and end-of-January 2022. This combined with school holidays in December and mid-February may also have contributed to a stagnation of RSV activity.

The year-round continuous RSV transmission pattern is distinctive not only from pre-pandemic winter epidemics in the Netherlands but also from neighboring countries such as Germany and France. These two countries returned to a winter epidemic in 2021 with low RSV activity during the summer of 2022. NPIs strictness and virus importation from external sources could explain these observed variations in RSV seasonality as these factors shape the level of population immunity debt [6, 9] (**Supplementary Figure 4C**). Furthermore, differences in surveillance and reporting strategies may explain the observed difference.

Our results suggest that although a shift in RSV seasonality may occur after implementation and subsequent relaxation of NPIs, RSV activity will most likely return to normal epidemic timing because herd immunity against RSV infection will return to pre-pandemic levels after two seasons of RSV exposure. To date, our model projection is aligned with the national RSV activity report from the National Institute for Public Health and the Environment (RIVM) [10].

Our study has several limitations. First, since not all participating hospitals used standard RSV testing, we may have underestimated RSV-related hospitalization rates. Some hospitals only used standard testing during the summer outbreak, which may have led to the underestimation of hospitalizations after the outbreak. Second, data collection was not standardized for each hospital. Although most hospitals shared prospective data, some used DBC codes or virology results to collect retrospective data. Third, we received consistent data from 34/46 hospitals for the entire study period. This may have led to underestimation of hospitalization rates. Lastly, our model was not calibrated to the observed hospitalization data in the Netherlands due to the short observational period. Instead, we simulated RSV epidemics using parameters from previous RSV models in the United States based on Dutch demographics. Although our simulations may not perfectly resemble the re-emergent RSV epidemics in the Netherlands, the flexible parameter ranges in our web-based app provide an opportunity for researchers from other countries to simulate re-emergent epidemics based on their local demographic and epidemiological characteristics.

In summary, we describe a distinct pattern of ongoing RSV transmission following an out-of-season RSV outbreak in the Netherlands after relaxation of COVID-19-related NPIs. The pattern is different from neighboring countries and may be partly explained by “immunity debt”, i.e. waning population immunity after a long period of low RSV exposure. Continuous monitoring of RSV seasonality using hospital-based data is essential to anticipate future RSV epidemics.

## Data Availability

Data and code to reproduce the simulation are available from GitHub (https://github.com/weinbergerlab/SPREAD.git).

## NOTES

## Acknowledgments

The authors thank the children and their families who participated in the study; Eline Bel, Katja Steenhuis, Marit de Bruijne, Kiora Russel, Nathalie Oldenburger, Kayleigh Vievermanns, Kim Bodaar, and Arda Yilmaz for their assistance in data collection; and all nurses, nurse practitioners, supporting staff, doctors and pediatricians for their substantial and ongoing effort for this study. Lastly, the authors thank the Dutch Working Group on Clinical Virology (NWKV) from the Dutch Society for Clinical Microbiology (NVMM) and all participating laboratories for providing the virological data from the weekly laboratory virological report.

## Disclaimer

The funder has no role in the design and conduct of the study; collection, management, analysis, and interpretation of the data; preparation, review, or approval of the manuscript; or decision to submit the manuscript for publication.

