## supplementary document for "Year-round RSV Transmission in the Netherlands Following the COVID-19 Pandemic - A Prospective Nationwide Observational and Modeling Study"

**SUPPLEMENTARY DATA**

**Supplementary Table 1**. Participating Hospitals.

| **Ziekenhuis** | **Type** | **Province** | **Targeted RSV testing** | **Age-stratified data collection** |
| --- | --- | --- | --- | --- |
| Admiraal de Ruyter Ziekenhuis | Regional | Zeeland | Standard | No |
| Albert Schweitzer | Regional | Zuid-Holland | Standard | No |
| Alrijne Ziekenhuis | Regional | Zuid-Holland | Standard | Yes |
| Amsterdam UMC | Academic | Noord-Holland | Standard | No |
| Antonius Ziekenhuis | Regional | Utrecht | Standard until June 2022 | No |
| Antonius Zorggroep Sneek | Regional | Friesland | Not standard | No |
| Beatrix Ziekenhuis | Regional | Zuid-Holland | Standard | No |
| Bernhoven ziekenhuis | Regional | Noord-Brabant | Standard | No |
| BovenIJ Ziekenhuis | Regional | Noord-Holland | Standard | No |
| Bravis Ziekenhuis | Regional | Noord-Brabant | Not standard | No |
| Catharina Ziekenhuis | Regional | Noord-Brabant | Not standard | No |
| Dijklander Ziekenhuis | Regional | Noord-Holland | Standard | No |
| Elizabeth-TweeSteden Ziekenhuis | Regional | Noord-Brabant | Standard | Yes |
| Elkerliek Ziekenhuis | Regional | Noord-Brabant | Standard until December 2021 | Yes |
| Erasmus Medisch Centrum | Academic | Zuid-Holland | Standard | No |
| Flevoziekenhuis | Regional | Flevoland | Standard | No |
| Franciscus Gasthuis & Vlietland | Regional | Zuid-Holland | Standard | Yes |
| Gelre Ziekenhuis | Regional | Gelderland | Not standard | No |
| Groene Hart Ziekenhuis | Regional | Zuid-Holland | Standard | No |
| Haaglanden Medisch Centrum | Regional | Zuid-Holland | Standard | No |
| Ijsselland Ziekenhuis | Regional | Zuid-Holland | Standard | No |
| Isala | Regional | Overijssel | Standard June 2021 - July 2022 | Yes |
| Leiden UMC | Academic | Zuid-Holland | Standard | No |
| Maastricht UMC | Academic | Limburg | Standard | No |
| Martini Ziekenhuis | Regional | Groningen | Standard | No |
| Maxima Medisch Centrum | Regional | Noord-Brabant | Standard | No |
| Meander Medisch Centrum | Regional | Utrecht | Standard until June 2022 | Yes |
| Medisch Centrum Leeuwarden | Regional | Friesland | Standard | No |
| Noordwest Ziekenhuisgroep | Regional | Noord-Holland | Not standard | No |
| OLVG | Regional | Noord-Holland | Standard | No |
| Radboud UMC | Academic | Gelderland | Standard | No |
| Rijnstate Ziekenhuis | Regional | Gelderland | Standard until June 2022 | Yes |
| Slingeland Ziekenhuis | Regional | Gelderland | Standard | No |
| Spaarne Gasthuis | Regional | Noord-Holland | Standard | Yes |
| Tergooi | Regional | Noord-Holland | Standard | No |
| Treant Zorggroep | Regional | Drenthe | Standard until June 2022 | No |
| UMC Groningen | Academic | Groningen | Standard | No |
| VieCuri Medisch Centrum | Regional | Limburg | Not standard | No |
| Wilhelmina Kinderziekenhuis Utrecht | Academic | Utrecht | Standard | Yes |
| Wilhelmina Ziekenhuis Assen | Regional | Drenthe | Standard | No |
| Zaans Medisch Centrum | Regional | Noord-Holland | Standard | No |
| Ziekenhuis Rivierenland | Regional | Gelderland | Standard until June 2022 | No |
| Ziekenhuis St Jansdal | Regional | Gelderland | Standard | Yes |
| Ziekenhuis Tjongerschans | Regional | Friesland | Standard | No |
| ZorgSaam | Regional | Zeeland | Standard | No |
| Zuyderland Medisch Centrum | Regional | Limburg | Standard until February 2022 | No |

Hospitals were invited to participate

**Supplementary Figure 1**. Distribution of Participating Hospitals in the Netherlands.
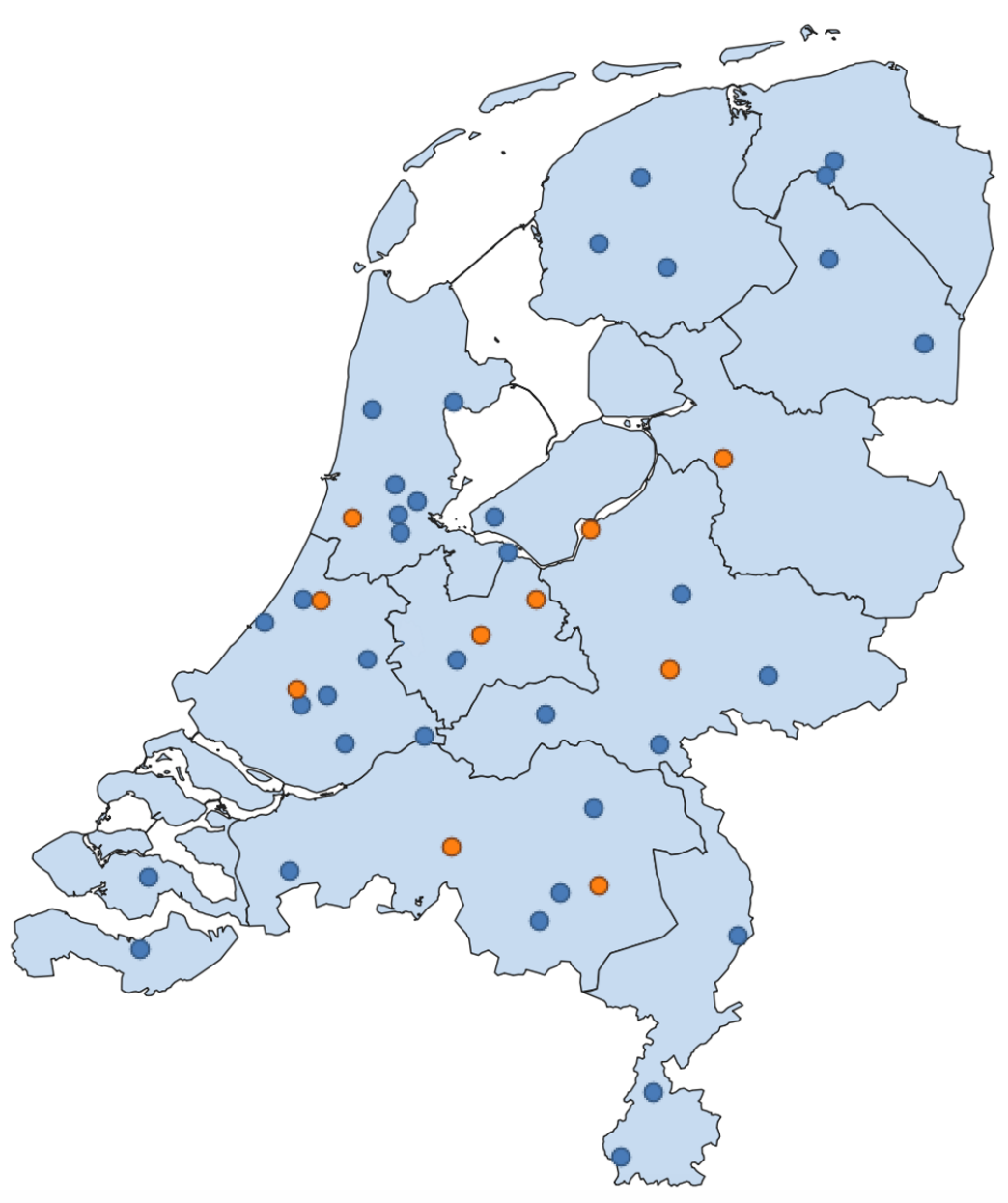

Blue, prospective data collection; orange, prospective data collection and age-stratified data collection.

**Supplementary Table 2.** Median Age Distribution of RSV-related Pediatric Hospital Admissions During 2018-2022.

|  | **Pre-COVID-19** | **Post-COVID-19** | | |  |
| --- | --- | --- | --- | --- | --- |
|  | **2018/2019 and 2019/2020 ‘regular’ winter season**  N=328 | **2021 summer outbreak (May-August)**  N=326 | **P-value** | **2021-2022 endemic phase (September 2021-August 2022)**  N=334 | **P-value*** |
| Median age [days, IQR] | 69 (35-176) | 161 (56-383) | <0.001 | 137 (54-281) | <0.001 |
| Proportion <6 months [N] | 76.5% (251) | 54.6% (178) | <0.001 | 60.5% (202) | <0.001 |

*2018-2020 ‘regular’ winter season vs 2021-2022 endemic transmission phase.

**Supplementary Table 3.** Median Age Distribution of RSV-related Pediatric Hospital Admissions During 2018-2022 excluding UMCU patients.

|  |  | **Pre-COVID-19** | **Post-COVID-19** | | |  |
| --- | --- | --- | --- | --- | --- | --- |
|  |  | **2018/2019 and 2019/2020 ‘regular’ winter season**  N=259 | **2021 summer outbreak (May-August)**  N=291 | **P-value** | **2021-2022 endemic phase (September 2021-August 2022)**  N=288 | **P-value*** |
|  | Median age [days, IQR] | 73 (36-187) | 165 (57-400) | <0.001 | 146 (59-303) | <0.001 |
| Proportion <6 months [N] | | 74.9% (194) | 53.4% (155) | <0.001 | 58.0% (167) | <0.001 |

*2018-2020 ‘regular’ winter season vs 2021-2022 endemic transmission phase.

**Supplementary Figure 2.** Median age of RSV hospitalization before and after the COVID-19 pandemic. The black line and values indicate the median age of RSV hospitalization (in days) varies before (including 2018-2019 and 2019-2020) and after the COVID-19 pandemic (2021-2022).

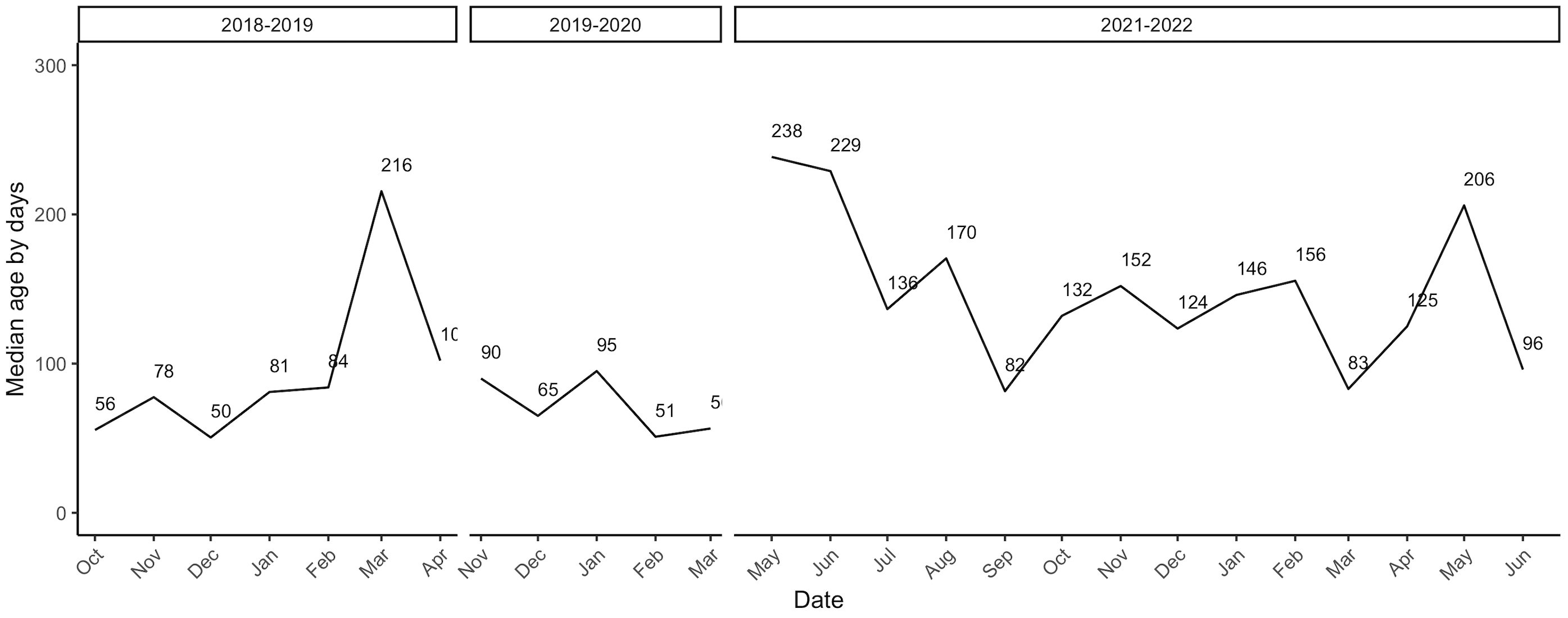

**Modeling Methodology**

*Transmission dynamic models*

We extended a previously published age-stratified RSV transmission model[1, 2]. The model assumes newborn infants are protected against RSV infections because they acquire neutralizing antibodies transplacentally from their mothers and/or have few contacts outside the household. With time, transplacentally acquired antibodies and cocooning effects wane, and infants become susceptible to infection. Following each infection, individuals gain partial immunity that lowers both their susceptibility to subsequent infections and the duration and infectiousness of subsequent infections (Supplementary Figure 3). The risk of lower respiratory disease depends on both the number of previous infections and age at infection in the model[1, 2].

Our previously published model, which assumed that partial immunity to reinfection did not wane, was not able to reproduce the pattern of endemic transmission that we observed during 2021-2022 in the Netherlands. Thus, we modified the model to allow for the partial immunity to wane without frequent exposure to RSV. We allowed partially individuals to revert to a more susceptible state after a period of months without reinfection.

We defined a seasonal age-specific force of infection that varies with time. The force of infection $\lambda_{a}\left( t \right)$ for age group $a$ and time *t* is defined as:

$$\lambda_{a}\left( t \right)=\left( 1+b_{1}cos\left( \frac{2\pi t-\phi}{12} \right) \right)\sum_{k} \beta_{a,k}(I_{1,k}\left( t \right)+\rho_{1}I_{2,k}\left( t \right)+\rho_{2}I_{3,k}\left( t \right){+\rho_{2}I_{4,k}\left( t \right))}/{N_{k}}\left( t \right)$$

It contains three major components: the seasonal transmissibility of RSV, the age-specific transmission parameter, and the transmissibility related to the number of infections. The seasonal dynamic of RSV is represented by $(1+b_{1}cos(\frac{2\pi t-\phi}{12}))$, where $b_{1}$is the amplitude of seasonality in transmission and *ϕ* is the timing of peak transmissibility. The transmission parameter $\beta_{a,k}$ is the product of the per capita probability of transmission given contact between an infectious and a susceptible individual (*q*) and contact rate between age group *k* and age group $a$ ($C_{a,k}$). We assume frequency-dependent age-specific contact patterns, which were obtained from a previous study that estimated the age-specific contact patterns for respiratory-spread infectious agents in the Netherlands [3]. The age-specific transmission parameter is multiplied by the number of infectious individuals of age *k* who have been infected one, two, three and four or more times at time $t$: ${(I}_{1,k}\left( t \right)+\rho_{1}I_{2,k}\left( t \right)+\rho_{2}I_{3,k}\left( t \right){+\rho_{2}I_{4,k}\left( t \right))}/{N_{k}}\left( t \right)$, where the relative infectiousness of second and subsequent infections are denoted as $\rho_{1}$ and $\rho_{2}$, respectively; the total population of age *k* at time $t$ is denoted as $N_{k}\left( t \right)$. We stratified the population into 21 age groups considering their risk of developing severe RSV disease and contact patterns. Infants younger than 12 months of age were divided into monthly age classes. The 1-4 years of age children were divided into yearly age classes. The remaining population was divided into 5 classes: 5–9 years, 10–19 years, 20–39 years, 40–59-years, and 60+ years of age.

The transmission dynamic process is linked to observed inpatient data. We assume that every infected individual has a probability $h_{i,a}$ of developing severe RSV disease that requires hospitalization, which depends on infection order *i* and age *a*, catchment area
$\eta$, and a fraction $\theta$ of RSV hospitalizations will be recorded in the inpatient datasets:

$$H_{a}\left( t \right)=\eta*\theta*(\lambda_{a}\left( t \right){(S}_{0,a}\left( t \right)h_{p,a}+{\sigma_{1}S}_{1,a}\left( t \right)h_{s,a}+{\sigma_{2}S}_{2,a}\left( t \right)h_{t,a}+{\sigma_{3}S}_{3,a}\left( t \right)h_{t,a}))$$

where $H_{a}\left( t \right)$ is the number of RSV hospitalizations in age group *a* at time *t* and $\lambda_{a}\left( t \right)$ is the force of infection that age group $a$ experiences at time $t$. The fully susceptible individuals of age *a* who have never been infected before are denoted as $S_{0,a}\left( t \right)$. The number of susceptible individuals of age *a* who have been infected one, two, and more times at time $t$ are denoted as $S_{1,a}\left( t \right), S_{2,a}\left( t \right),$ and $S_{3,a}\left( t \right)$, respectively; $\sigma_{1}, \sigma_{2},$ and $\sigma_{3}$ represent the reduced susceptibility to RSV infection following the first, second, and more infections due to the partial immunity gained after each infection. $h_{p,a}$, $h_{s,a}$ and $h_{t,a}$ are the proportion of the first, second, and subsequent infections in age group $a$ that require hospitalizations, respectively.

Since this study only recorded one typical seasonal RSV epidemic in children under 2 years of age before COVID-19 in the Netherlands, we did not fit the model to inpatient dataset to estimate the exact transmission parameters. Instead, we performed a simulation study with parameters drawn from reasonable ranges, which were informed by the published literature or based on estimates of the fitted parameters for models describing 9 years of inpatient data in the United States (supplementary table 4). We simulated the model for 7 years (July 2018 to June 2025) following a burn-in period of 19 years.

We explored four scenarios corresponding to hypotheses about the unusual seasonal RSV patterns following the COVID-19 pandemic. The four scenarios that we evaluated were (supplementary table 5):

(1) moderate level of NPIs and low level of virus importation, no waning immunity;

(2) moderate level of NPIs and low level of virus importation, with waning immunity;

(3) moderate level of NPIs and high level of virus importation, with waning immunity;

(4) strict NPIs but high level of virus importation, with waning immunity.

We did not explore the impact of climate variables because the temperature, vapor pressure, precipitation, and evapotranspiration did not change significantly in 2021 and 2022.

**Supplementary Figure 3.** Model structure.

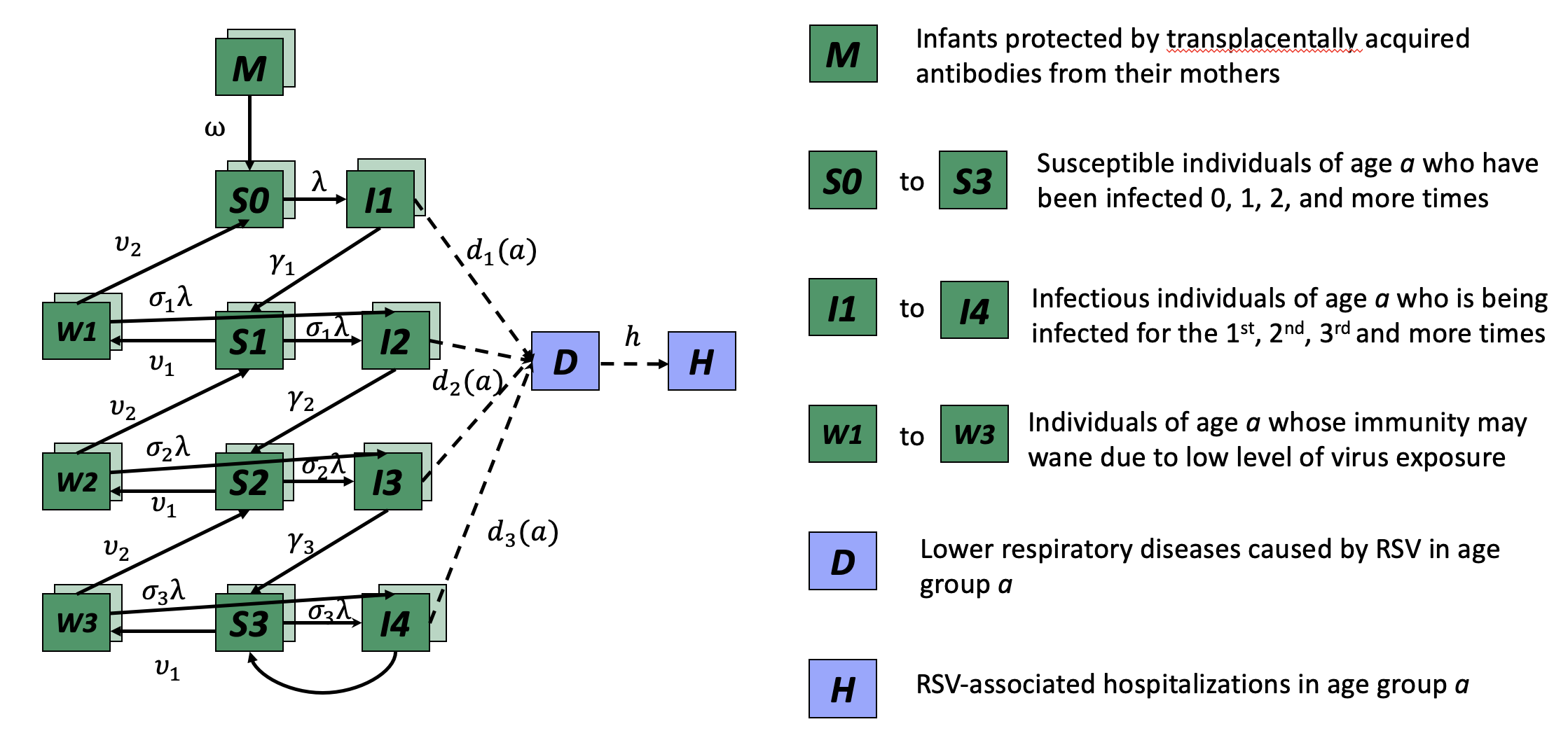

The green boxes represent infection states in the model, while purple boxes represent diseased states (RSV lower respiratory illness, D, and RSV hospitalizations, H).

**Supplementary Table 4. Model parameters.**

| **Parameter description** | **Symbol** | **Parameter value mean (range of value)** | **Refer-ence** | **Note** |
| --- | --- | --- | --- | --- |
| **Transmission dynamic models** | | | | |
| Duration of infectiousness | | | | |
| First infection | $1/\gamma_{1}$ | 10 (3, 28) days | [4-6] |  |
| Second infection | $1/\gamma_{2}$ | 7 (3, 8) days |  |  |
| Subsequent infection | ${1/\gamma}_{3}$ | 5 (1, 6) days |  |  |
| Relative risk of infection following | | | | |
| First infection | $\sigma_{1}$ | 0.76 (0.6, 1) | [7-10] |  |
| Second infection | $\sigma_{2}$ | 0.6 (0.4, 0.8) |  |  |
| Subsequent infection | $\sigma_{3}$ | 0.4 (0.2, 0.6) |  |  |
| Relative infectiousness | | | | |
| Second infections | $\rho_{1}$ | 0.75 (0.5, 1) | [7, 8, 11] |  |
| Subsequent infections | $\rho_{2}$ | 0.51 (0.25, 0.75) |  |  |
| Proportion of RSV infections leading to hospitalization | | | | |
| First infection |  |  | [11-17] | The probability of hospitalization given infection was estimated as the product of the probability of hospitalization given lower respiratory tract infections, the probability of lower respiratory tract infections given symptomatic infection (*I_S_*), and the probability of symptoms given infection: $\Pr\left( hosp \vert I \right)=\Pr\left( hosp \vert LRI \right)*\Pr\left( LRI \vert I_{s} \right)*\Pr\left( I_{s} \vert I \right)$  We estimated the age-specific probability by fitting polynomial regressions to reported aggregate probabilities for 3-month or 6-month age groups |
| 0-2 months old | $h_{p,0-2}$ | 0.072 |  |  |
| 3-5 months old | $h_{p,3-5}$ | 0.0312 |  |  |
| 6-8 months old | $h_{p,6-8}$ | 0.016 |  |  |
| 9-11 months old | $h_{p,8-9}$ | 0.012 |  |  |
| 1 year old | $h_{p,1y}$ | 0.0096 |  |  |
| 2-4 years old | $h_{p,2-4y}$ | 0.007 |  |  |
| $\geq5$ years old | $h_{p,5+y}$ | 0.001 |  |  |
| Second infection | $h_{s,a}$ | 0.4*$h_{p,a}$ |  |  |
| Third+ infection | $h_{t,a<40}$ | 0 |  |  |
| 40-59 years old | $h_{t,40-59}$ | 0.00001 | [8] |  |
| 60+ years old | $h_{t,60+}$ | 0.00004 |  |  |
| Duration of boosted immunity (days) | $1/\upsilon_{1}$ | 180 (90, 40000) days | [18] | Upper bound equals no waning during lifetime |
| Duration of partially boosted immunity (days): | $1/\upsilon_{2}$ | 200 (90, 40000) days | [18] |  |
| Transmission parameter* | $q$ | 8.5 $(7, 11)$ | [1, 2] |  |
| Amplitude of seasonality | $\alpha$ | 0.13 (0.10,0.30) | [1, 2] |  |
| Timing of seasonality | $\phi$ | 3.56 (0,2π) |  |  |
| Reporting fraction^+^ | $\theta$ | 0.45 (0.2,0.8) | [1, 2] |  |
| Catchment area | $\eta$ | 0.5 (0,1) |  |  |

*The basic reproductive number (*R*_0_) was estimated from $R_{0}=\frac{det(\beta_{a,k})}{\gamma_{1}}=\frac{det({qC}_{a,k})}{\gamma_{1}},$ using the next-generation matrix method; the transmission parameter *q* was informed by previous study that fitted to the U.S. inpatient data, $C_{a,k}$ is the contact matrix scaled by the proportion of the population within each age class.

^+^ Reporting fraction is a product of the probability that children got tested for the pathogens causing the infection, the probability that the result is positive given RSV infection, and the reporting and recording rate of doctors when they receive positive RSV tests.

**Supplementary Table 5. Scenario parameters.**

| **Scenario** | **Scenario description** | **Parameter value** |
| --- | --- | --- |
| 1 | moderate level of NPIs and low level of virus importation, no waning immunity | Duration of constant NPIs: 11 months  Duration of gradual NPIs relaxation: 3 months  Percent decrease in transmission: 28%  Virus importation: 1/1000000 travelers per month  Duration of waning immunity: 12 months  Approximate R0: 10  Amplitude of seasonality: 0.3  Phase of seasonality: 3.41 |
| 2 | moderate level of NPIs and low level of virus importation, with waning immunity | Duration of constant NPIs: 11 months  Duration of gradual NPIs relaxation: 3 months  Percent decrease in transmission: 28%  Virus importation: 1/1000000 travelers per month  Duration of waning immunity: 12 months  Approximate R0: 8.5  Amplitude of seasonality: 0.13  Phase of seasonality: 3.56 |
| 3 | moderate level of NPIs and high level of virus importation, with waning immunity | Duration of constant NPIs: 11 months  Duration of gradual NPIs relaxation: 3 months  Percent decrease in transmission: 28%  Virus importation: 50/1000000 travelers per month  Duration of waning immunity: 12 months  Approximate R0: 8.5  Amplitude of seasonality: 0.13  Phase of seasonality: 3.56 |
| 4 | strict NPIs but high level of virus importation, with waning immunity | Duration of constant NPIs: 14 months  Duration of gradual NPIs relaxation: 3 months  Percent decrease in transmission: 28 %  Virus importation: 50/1000000 travelers per month  Duration of waning immunity: 12 months  Approximate R0: 8.5  Amplitude of seasonality: 0.13  Phase of seasonality: 3.56 |

**Supplementary Figure 4**. **The simulated number of monthly respiratory syncytial virus (RSV) hospitalizations in children <2 years of age under alternative scenarios, 2018–2025.** The projected RSV hospitalizations under alternative scenarios 1, 3, and 4 after the interruption of COVID-19-related measures over time are plotted for July 2018 to June 2025. The green line corresponds to the total RSV hospitalizations in children under 2 years of age. The orange line corresponds to the RSV hospitalizations in infants under 6 months of age. The purple line corresponds to the RSV hospitalizations in children 1 year of age. The shaded area corresponds to the period of the SPREAD study, May 2021 - August 2022. (A) Scenario 1: moderate level of NPIs and low level of virus importation, no waning immunity (B) Scenario 3: moderate level of NPIs and high level of virus importation, with waning immunity (C) Scenario 4: strict NPIs but high level of virus importation, with waning immunity. This scenario is similar to the observation in France and Germany with a few parameter adjustments. We assumed that COVID-19-related public health measures started at the end of March 2020 and gradually relaxed since the end of June 2021 over a 3-month period and that virus importation is 30 per 1 million travelers per month [19].

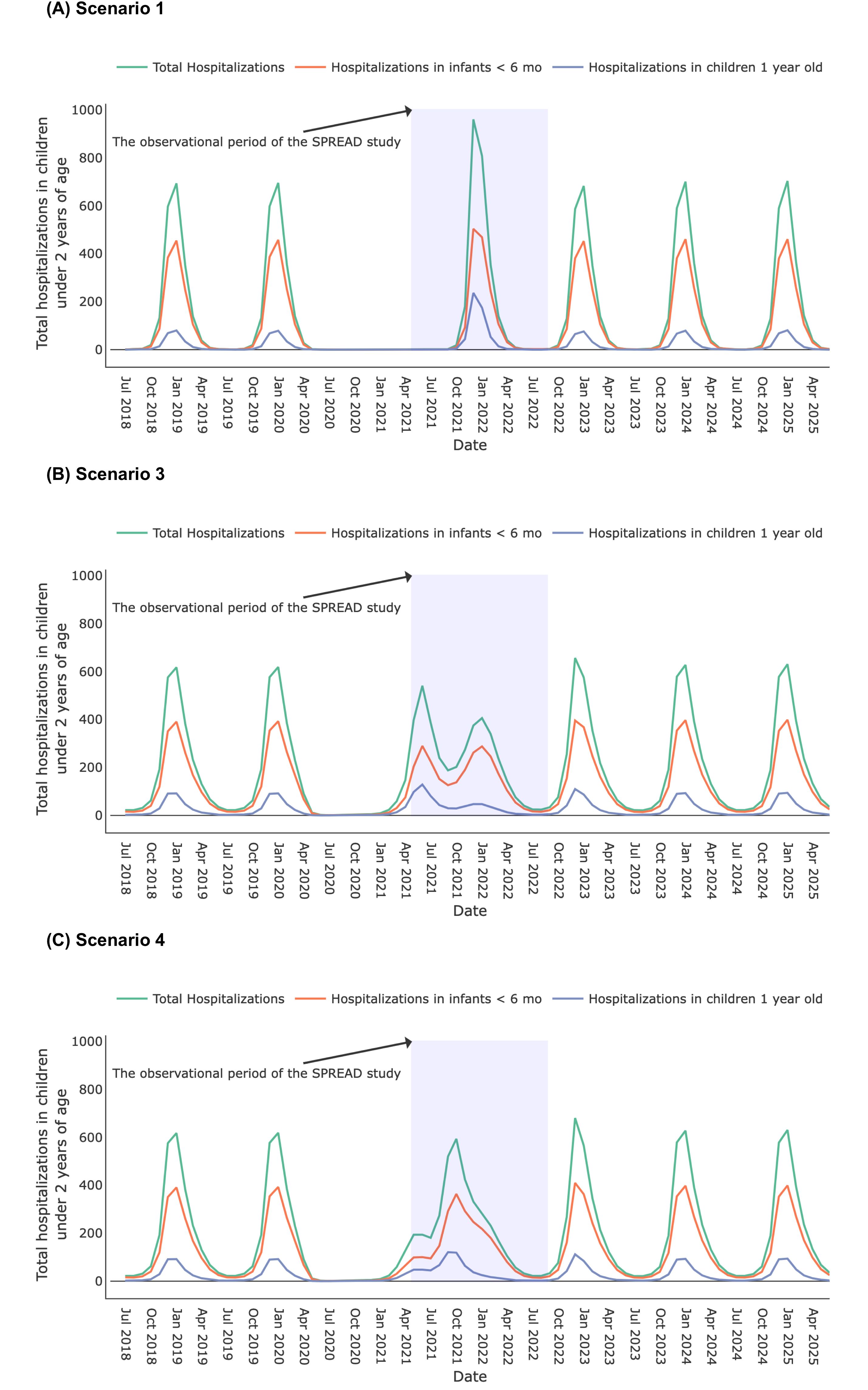

**Supplementary Figure 5**. **The** **projected total number of individuals in each of the susceptible compartments under Scenario 2, 2018–2025.** The green line corresponds to the total number of individuals that have low susceptibility to RSV infection. The pink line corresponds to the total number of individuals that have moderate susceptibility to RSV infection. The purple line corresponds to the total number of individuals that have high susceptibility to RSV infection. The orange line corresponds to the total number of individuals that never had RSV infection before.

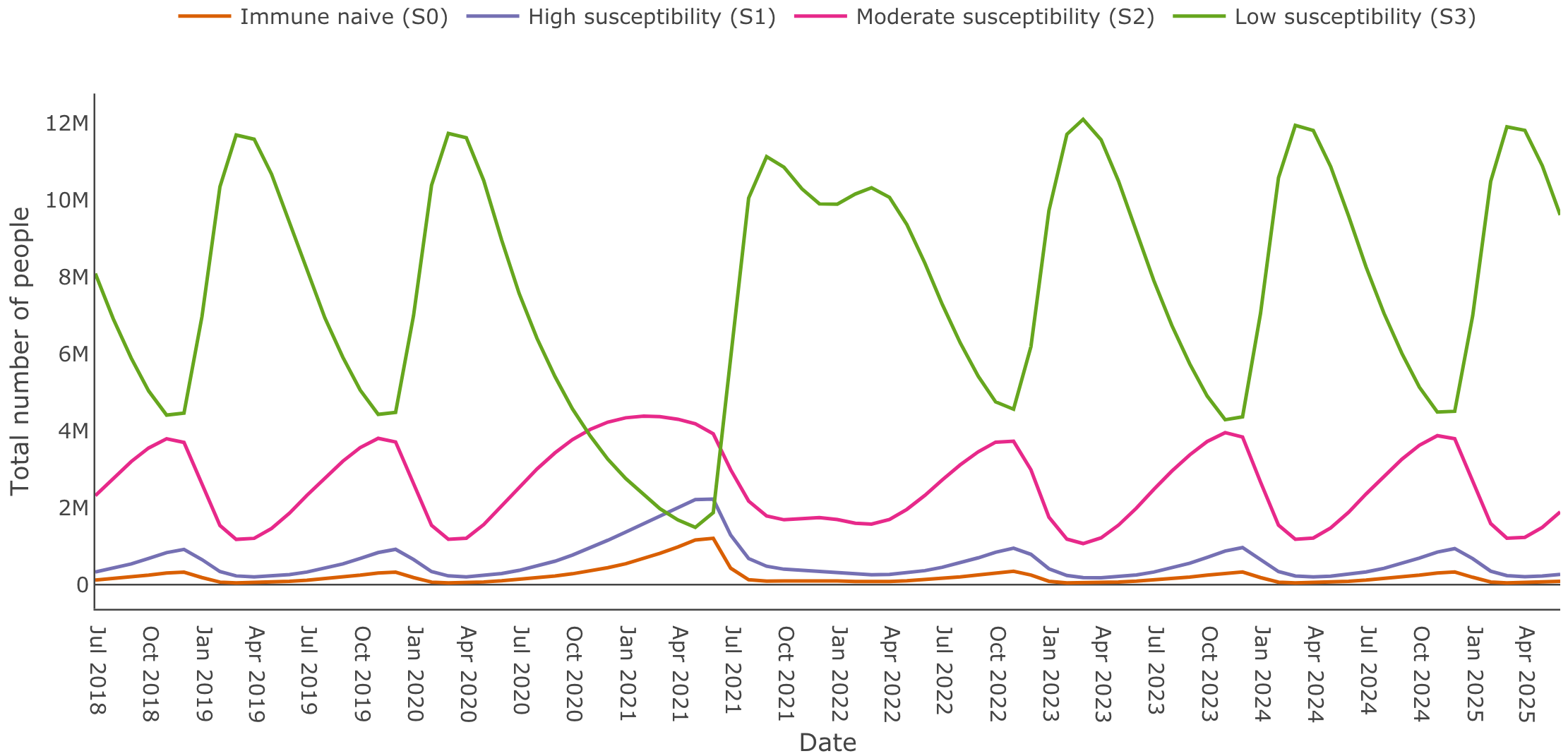

19.	ECDC. Data on country response measures to COVID-19. Available at: <https://www.ecdc.europa.eu/en/publications-data/download-data-response-measures-covid-19>.
